# Classification of scenarios based on the determinants of childhood vaccination coverage in Brazil

**DOI:** 10.64898/2026.07.17.26358349

**Authors:** Isaac Negretto Schrarstzhaupt, Francieli Fontana Sutile Tardetti Fantinato, Lely Stella Guzman-Barrera, Fredi Alexander Diaz-Quijano

## Abstract

**Background:** Vaccination coverage in Brazil declined between 2015 and 2022, followed by a recovery in 2023 and 2024. Given this instability, driven by multiple determinants, we sought to identify factors associated with childhood vaccination and group Brazilian municipalities into scenarios that could guide interventions to improve coverage.

**Methods:** In this ecological study of 5,274 Brazilian municipalities, we assessed the rate of children under 1 year unvaccinated with the third dose of the Inactivated Poliovirus Vaccine (IPV) in 2024, chosen for its high correlation with other tracer vaccines such as Diphtheria, Tetanus and Pertussis (DTP) and Measles, Mumps and Rubella (MMR). We applied a hierarchical model with three levels of determinants (socioeconomic, health service structure, and operational), using Poisson regression to estimate standardized Rate Ratios (RR), followed by a K-means cluster analysis to identify municipal profiles.

**Results:** Several determinants were associated with higher rates of unvaccinated children, most notably the proportion of the population not covered by Community Health Workers (CHW) (RR = 1.15; 95% CI 1.14-1.15) and inequality measured by the Gini Index (RR = 1.33; 95% CI 1.32-1.34). We identified six municipal profiles that differed in tracer-vaccine coverage up to four years of age and in the composite Vaccination Needs Index (VNI).

**Conclusion:** Multiple socioeconomic, structural and operational factors were associated with unvaccinated rates, highlighting the relevance of healthcare service organization even in favorable social contexts. Classifying municipalities into risk profiles may support more targeted interventions aligned with local needs.

## INTRODUCTION

Vaccination is recognized as one of the most successful and cost-effective public health interventions in history [1,2]. However, Brazil recorded a downward trend in vaccination coverage between 2015 and 2022, a situation that increased the risk of resurgence of already controlled vaccine-preventable diseases [3,4]. The possible causes of this decline are multifactorial and complex [5–7].

The strong influence of socioeconomic determinants, such as income inequality and the level of human development, on access and adherence to vaccination is already established in the scientific literature [8–12]. Although these factors are important for understanding the overall picture, they are slow to be changed in the short term by health managers. In addition, there is an important knowledge gap regarding the impact of more proximal determinants, directly linked to the infrastructure and daily operation of services, on the recovery and maintenance of vaccination coverage.

Understanding the impact of these operational factors is essential to support the definition of agile interventions at the local level [13]. In this study we assessed, from a hierarchical perspective, contextual factors associated with the lack of a dose of a tracer vaccine of the basic childhood schedule in Brazilian municipalities. Then, based on these factors, we identified groups of municipalities with similar profiles. This classification may help guide microplanning strategies and interventions more relevant to local needs [14].

## MATERIALS AND METHODS

This was a nationwide ecological study using secondary data from public sources, in which the analysis units were the Brazilian municipalities [15–17]. During the exploratory analysis of the variables we observed that the municipalities of São Paulo and Rio de Janeiro exerted a disproportionate influence on the regression models because of their larger population scale, and were therefore excluded from the analysis [18]. The final sample consisted of 5,568 municipalities, of which 5,274 had complete data and were used in the regression and cluster analyses (described below), while the remaining 294 municipalities, with incomplete data, were not submitted to cluster analysis and were kept as a separate incomplete data group (Figure 1). This group was used only for the validation stage regarding its vaccination indicators.

**Figure 1:**
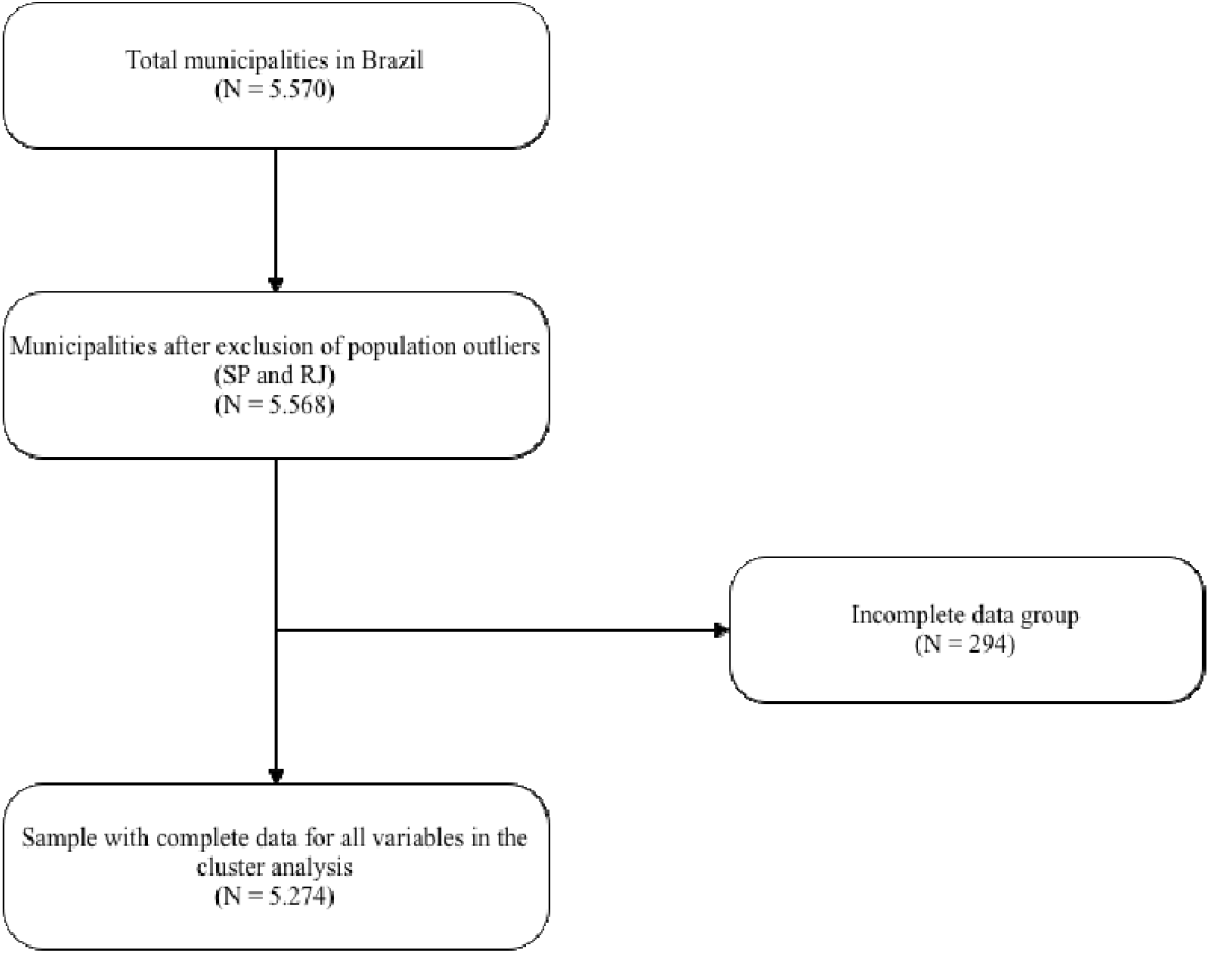
Flowchart of sample formation for the regression and cluster analyses.

To integrate the different databases, the Brazilian Institute of Geography and Statistics (IBGE) code of each municipality was used as the primary aggregation key. The data analyzed in this study were obtained within the scope of epidemiological surveillance activities, meaning the study was exempt from review by a Research Ethics Committee, in accordance with Resolution 466/2012 of the Brazilian National Health Council.

To select the outcome, we considered tracer vaccines that make up the national immunization schedule for children under 1 year of age, including DTP, IPV and MMR [19]. Due to the high statistical correlation observed among these vaccines, we chose the third dose of IPV in 2024. This vaccine showed an intermediate coverage frequency, which we considered would be more informative of the variability in coverage across municipalities (Supplementary Table S1) [20,21]. In addition, coverage of this vaccine is considered one of the most sensitive indicators monitored by the World Health Organization (WHO) and the National Immunization Program (PNI) of the Brazilian Ministry of Health to assess the risk of outbreaks and to sustain the mission of eliminating all wild polioviruses toward global eradication [22–26].

To guide the analyses, we adopted a hierarchical conceptual model that organizes the variables into three levels:

- **Distal level: Socioeconomic and demographic variables**. We included variables such as the Gini Index, per capita income, the Municipal Human Development Index (MHDI), life expectancy, rural population, literacy, and child labor. Data were extracted from TABNET, the Atlas of Human Development in Brazil, and the IBGE, mostly referring to 2010, the most recent period available at the time of this study [15,17,27–29].
- **Intermediate level: Health service structure variables**. This level included indicators such as Primary Health Care (PHC) coverage, information technology facilities, the number of computers, and the availability of internet in vaccination rooms [30].
- **Proximal level: Operational variables**. These variables, as well as the intermediate level ones, were extracted from a survey voluntarily completed by the teams of primary care units (Basic Health Units, UBS) within the National Program for Improving Access and Quality of Primary Care (PMAQ), conducted in 2015, and reflect the daily operation of the services [30]. Examples include: “Does the UBS carry out active search for children with delayed vaccination?”, “Does the UBS have a dedicated vaccination room?” and “Does the UBS open during lunchtime?” (Supplementary material).

### Data analysis

Statistical analysis was performed using Stata 18.0. We first sought the functional form of the continuous independent variables in relation to the outcome. As an example, we highlight the relationship between PHC coverage in 2023 and the number of unvaccinated children with the third dose of IPV in 2024 (Figure 2).

**Figure 2:**
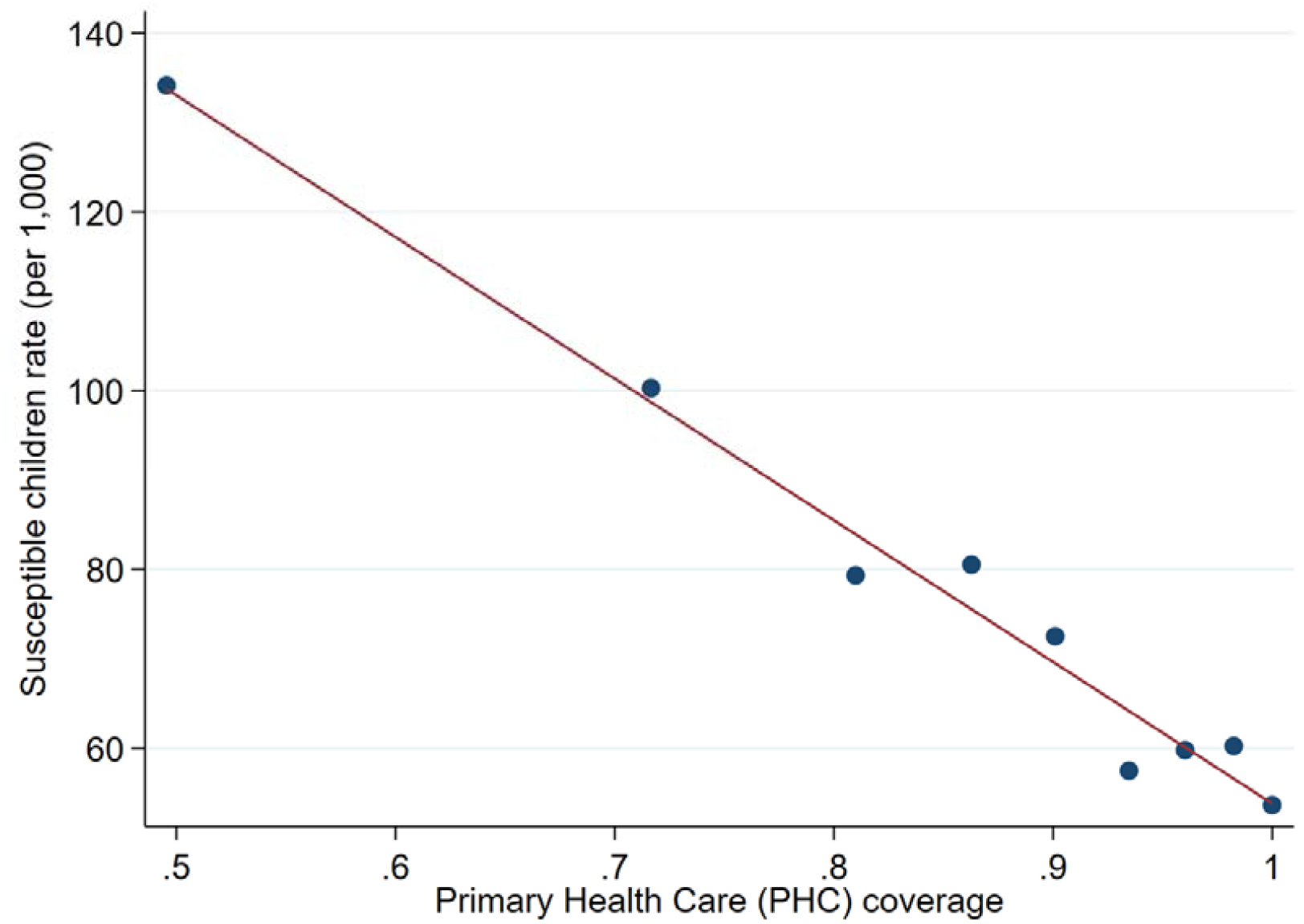
Linear relationship between Primary Health Care (PHC) coverage and the rate of children not vaccinated with the third dose of IPV (per 1,000), grouped by deciles, in Brazilian municipalities.

The PMAQ variables (originally with categorical “Yes/No” responses) were transformed, for each municipality, into the ratio between the teams that answered affirmatively and the total number of units that answered that question. All independent variables (socioeconomic, structural, and operational) were then standardized (transformed into Z-Scores) to improve the comparability of the association measures across determinants.

Poisson regression was used, with the number of susceptible children (under 1 year of age) as the dependent variable and the target population under 1 year of age in 2024 as the offset. As a measure of association, we calculated the Rate Ratio (RR) per one standard-deviation increase in the independent variable, with 95% confidence intervals.

### Interpretation of associations and cluster analysis

The associations were interpreted based on a hierarchical conceptual framework [31]. The first model (distal) estimated the total effects of the socioeconomic variables on the outcome, assuming independence of mechanisms. In a second model (intermediate), we evaluated structural health variables, adjusted for the previous level. Finally, the last model assessed the effect of the operational variables (proximal), adjusted for the distal and intermediate determinants. In other words, as illustrated in the Directed Acyclic Graph (DAG, Figure 3), when variables from all levels are included, only those of the proximal variables would be interpretable as total effects. The full hierarchical models, with all variables of each level, are presented in Supplementary Table S2.

**Figure 3:**
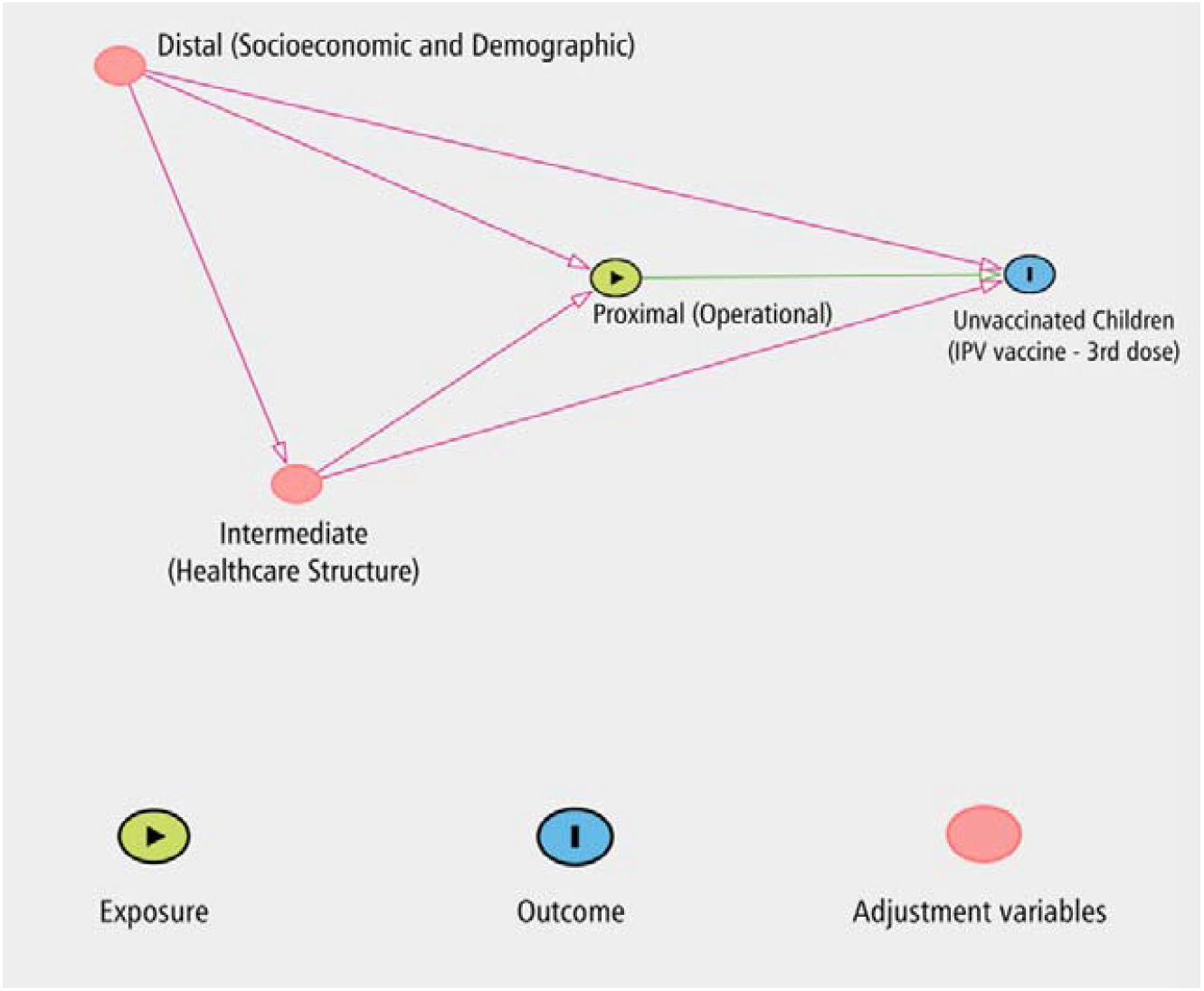
Directed Acyclic Graph (DAG) showing the relationship between the three hierarchical levels.

We anticipated that some associations could show counterintuitive results due to potential reverse causality phenomena. For example, a higher frequency of active search for susceptible children may occur precisely as an emergency response to a prior situation of poorer vaccination coverage in the municipality. To mitigate these phenomena, the subsequent step of defining intervention scenarios (clusters) included only the variables whose association was both statistically significant and considered intuitive by the authors.

Those variables were then used to identify clusters using the K-means algorithm, following a methodological approach previously described for defining scenarios in public health [32,33]. The final choice of six clusters (k=6) was based on the predictive capacity of the model, whose Pseudo R^2^ (7.5%) represented a gain compared with the five-cluster solution (4.4%) [34]. The clusters were named based on the authors’ interpretation, according to the distributions of the determinants identified in the analysis.

To assess the consistency of this classification, we compared the profiles with respect to measures of vaccination status (not included in the cluster analysis). Among these measures, we considered the cumulative vaccination coverage of three tracer doses (third dose of DTP, third dose of IPV and second dose of MMR) of the 0-4-year-old cohort in 2024. Because it includes children vaccinated over the past years, encompassing the period of declining coverage (2015-2022), this cohort tends to show lower values than those observed among children under 1 year of age in 2024. In addition, we calculated the Vaccination Needs Index (VNI), which integrates absolute and relative indicators of the tracer vaccines of the Brazilian schedule. Higher VNI values indicate greater vaccination needs, reflecting lower coverage and/or a larger susceptible population [21].

## RESULTS

In 2024, the mean coverage for the third dose of IPV among children under 1 year of age, across the 5,274 municipalities with complete data (excluding São Paulo and Rio de Janeiro), was 93.6%. In aggregate, 214,709 children under one year of age remained susceptible (without the complete vaccination schedule for their age), corresponding to an overall (population-weighted) coverage of 89%. This value is lower than the 93.6% because the latter is the unweighted mean of municipal coverages, whereas the population-weighted estimate accounts for the number of children in each municipality. Since several more populous municipalities had lower coverage, the weighted figure is lower.

Considering the distal-level variables, those negatively associated with the rate of unvaccinated children were the MHDI (RR = 0.61; 95% CI 0.60-0.62), life expectancy (RR = 0.93; 95% CI 0.92-0.94), per capita income (RR = 0.98; 95% CI 0.98-0.99) and rural population (RR = 0.71; 95% CI 0.71-0.72). The Gini Index, in turn, was positively associated with the outcome (RR = 1.33; 95% CI 1.32-1.34). Moreover, there were differences among the country’s regions, with a higher rate of unvaccinated children in the North region (Figure 4A). Conversely, associations that we considered counterintuitive were observed for literacy rates (RR = 1.73; 95% CI 1.71 – 1.75) and child labor (RR = 0.94; 95% CI 0.93-0.95) (Table 1).

**Table 1:** Poisson regression models with factors associated with the rate of unvaccinated children.

| Standardized (Z-Scores) variables | Mean (SD) <sup>a</sup> | RR (95% CI) <sup>b</sup> |
| --- | --- | --- |
| *Distal level (Socioeconomic) <sup>d</sup>* |  |  |
| Gini index | 0.50 (0.07) | 1.33 (1.32-1.34) |
| Life expectancy, in years | 73.02 (2.68) | 0.93 (0.92-0.94) |
| MDHI | 0.66 (0.07) | 0.61 (0.60-0.62) |
| Rural population | 31.1 (19.9) | 0.71 (0.71-0.72) |
| PCI (R\$) <sup>c</sup> | 474.80 (230.61) | 0.98 (0.98- 0.99) |
| Literacy rate | 88 (7.6) | 1.73 (1.71-1.75) |
| Child labor | 13 (8.3) | 0.94 (0.93-0.95) |
| *Intermediate level (Health services structure) <sup>d</sup>* |  |  |
| PHC coverage | 93.8 (12.3) | 0.82 (0.82-0.82) |
| Internet in vaccine room | 80.5 (31.6) | 0.81 (0.81-0.82) |
| Computers in vaccine room | 3.14 (5.33) | 1.04 (1.04-1.04) |
| *Proximal level (Operational) <sup>d</sup>* |  |  |
| Always offers vaccine | 78.9 (31) | 1.05 (1.04-1.05) |
| Opens in lunch hours | 43.5 (41.1) | 0.98 (0.97-0.99) |
| Does active search | 97.1 (10.8) | 1.02 (1.01-1.02) |
| Does not do active search | 1.4 (7.7) | 1.02 (1.01-1.03) |
| Population without CHWs coverage | 30.4 (34.5) | 1.15 (1.14-1.15) |

Notes:
a) SD: Standard Deviation. Values are presented on their original scale for better understanding.
b) RR: Rate Ratio. 95% CI: 95% Confidence Interval. Standardized variables (Z-Scores) for the Poisson analysis. The RR represents the relative change in the rate of susceptible children for each 1-standard-deviation increase in the independent variable, adjusted for the other variables. p-value of all associations presented in this table was < 0.001.
c) Values expressed in Brazilian Real (BRL). At the time of socioeconomic data collection (2010), the average exchange rate was US\$ 1.00 = BRL 1.76 [35].
d) The measures of association reflect the hierarchical approach, presenting total effects for each level, with levels 2 and 3 adjusted by its predecessors.

**Figure 4:**
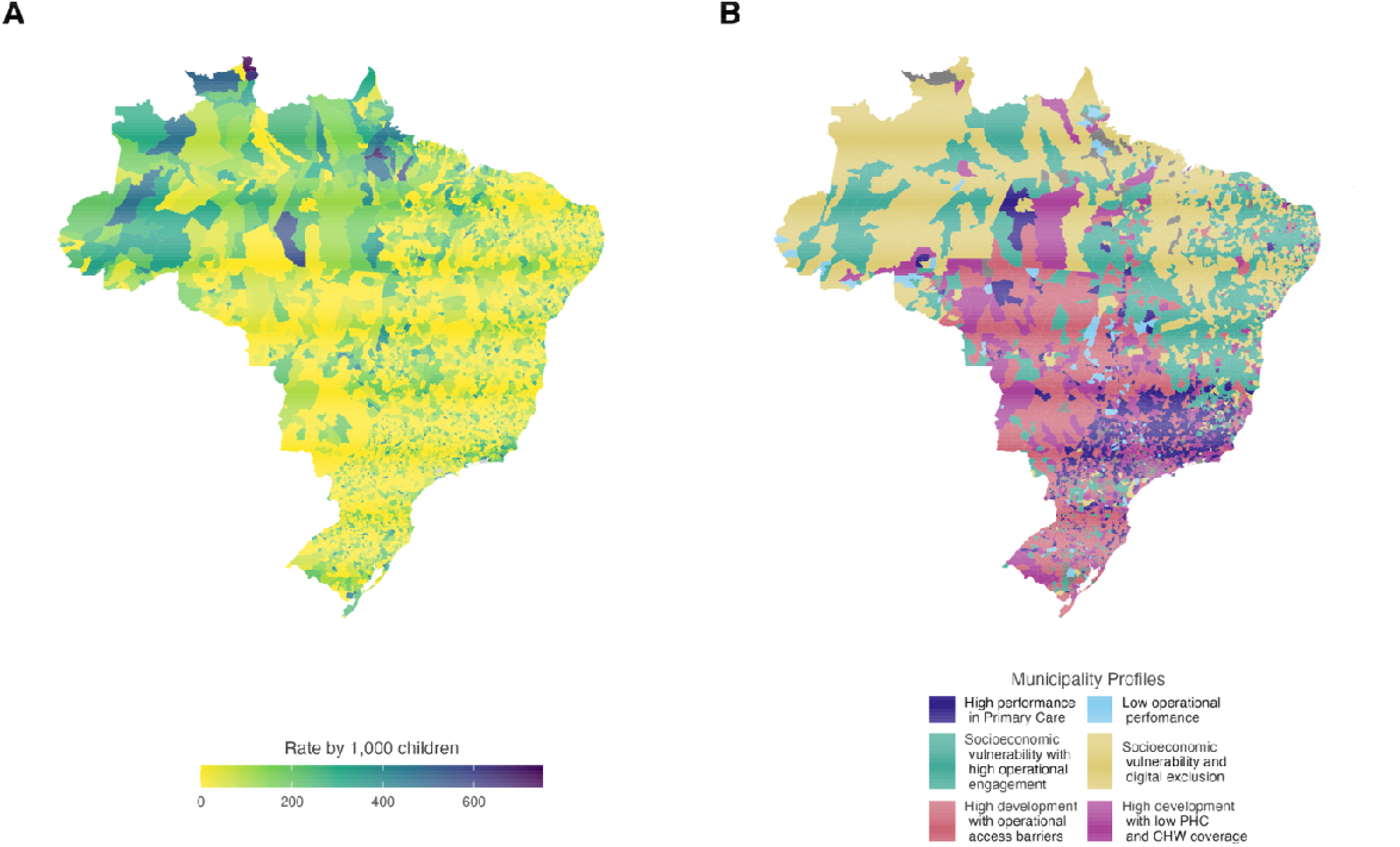
Spatial distribution of the rate of unvaccinated children for the third dose of IPV (4A) and the identified municipal clusters (4B) in Brazil.

At the intermediate level, the structural variables presented protective and risk associations with the rate of unvaccinated children. After adjustment for the preceding block, PHC coverage (RR = 0.82; 95% CI 0.82-0.82) and the presence of internet in the units (RR = 0.81; 95% CI 0.81-0.82) were strong protective factors. We considered the positive association between the presence of computers in the vaccination room and the outcome to be counterintuitive (RR = 1.04; 95% CI 1.04-1.04).

The proximal-level variables, adjusted for the previous levels, showed the strongest associations with the outcome (Table 1). Notable findings were the risk associated with the population not covered by Community Health Workers (CHW) (RR = 1.15; 95% CI 1.14-1.15) and the protective effect of keeping the UBS open during lunchtime (RR = 0.98; 95% CI 0.97-0.99). Not performing active search (a strategy for screening and recovering children with delayed schedules in the territory) was also positively associated (RR = 1.02; 95% CI 1.01-1.03). In contrast, the regular supply of vaccines and the active search for defaulters in the routine program showed counterintuitive associations (RR = 1.05; 95% CI 1.04-1.05 and RR = 1.02; 95% CI 1.01-1.02, respectively).

### Characterization of municipal profiles

The cluster analysis characterized six distinct municipal profiles in Brazil (Table 2). The profile we named “High performance in Primary Care” (20.9% of municipalities) showed the most egalitarian scenario (lowest Gini) and good management indicators, such as keeping units open during lunchtime. Similarly, the “Socioeconomic vulnerability with high operational engagement” profile (26.4%), despite its lower socioeconomic level, showed high PHC coverage, internet access in vaccination rooms, and a low proportion of the population not covered by CHW.

**Table 2:** Characterization of Municipality Profiles based on Hierarchical Determinants ^a^.

|  | High performance in Primary Care (N = 1,101) | Socioeconomic vulnerability with high operational engagement (N = 1,394) | Low operational performance (N = 100) | Socioeconomic vulnerability and digital exclusion (N = 955) | High development with operational access barriers (N = 1,188) | High development with low PHC and CHW coverage (N = 536) |
| --- | --- | --- | --- | --- | --- | --- |
| *Level 1: Socioeconomic* |  |  |  |  |  |  |
| MHDI <sup>b</sup> | 0.700 | 0.598 | 0.679 | 0.583 | 0.708 | 0.740 |
| Life expectancy, in years <sup>b</sup> | 74.75 | 70.88 | 73.99 | 70.35 | 74.91 | 75.43 |
| Gini index <sup>b</sup> | 0.458 | 0.526 | 0.499 | 0.554 | 0.476 | 0.510 |
| PCI (R\$) <sup>b</sup> | 512.12 | 288.39 | 544.29 | 260.32 | 639.08 | 764.74 |
| Rural population (%) <sup>b</sup> | 23 | 40.5% | 27.5% | 43.5% | 28.6% | 7.4% |
| *Level 2: Health services structure* |  |  |  |  |  |  |
| PHC coverage (%) <sup>b</sup> | 98.1 | 85.4 | 88.8 | 90.9 | 98 | 77.5 |
| Internet in vaccine room (%) <sup>b</sup> | 95.5 | 90.8 | 88.3 | 24.4 | 96 | 87.1 |
| Computers in vaccine room <sup>b</sup> | 1.78 | 2.92 | 1.28 | 2.66 | 2.07 | 10.01 |
| *Level 3: Operational* |  |  |  |  |  |  |
| Open in lunch hours (%) <sup>b, c</sup> | 92.7 | 37.8 | 36.9 | 34.6 | 5.8 | 57.9 |
| Not doing active search (%) <sup>b, c</sup> | 0.3 | 0.2 | 48.4 | 0.8 | 0.3 | 1.4 |
| Population without CHWs coverage (%) <sup>b</sup> | 20.6 | 21.5 | 55.5 | 32.6 | 30.3 | 64.9 |

*Notes:*
a) Variables with counterintuitive results from the regression analysis were not used in the cluster analysis.
b) average values for the municipalities in each profile (cluster).
c) percentage of basic health units that answered affirmatively in the PMAQ survey among those that responded to the question, averaged across municipalities in each profile.

In contrast, “Low operational performance” grouped a small number of municipalities (1.9%) with favorable socioeconomic indicators, such as a mean MHDI of 0.679. This profile showed critical operational deficiencies, with 48.4% of the teams responding to the survey not performing active search for defaulters. Additionally, this profile recorded the second-highest proportion of the population not covered by CHW (55.5%).

Profiles marked by high vulnerability were also characterized: “Socioeconomic vulnerability and digital exclusion” (18.1%) grouped the municipalities with the lowest mean MHDI (0.583) and mean per capita income (R$ 260.32). Digital infrastructure was the most limited, with only 24% of their health units responding to the PMAQ survey having internet access in vaccination rooms. In the validation stage, this profile showed low vaccination coverage.

The “High development with operational access barriers” profile (22.5%) brought together municipalities with good service structure but was characterized by access limitations, evidenced by the lowest proportion of units open during lunchtime. In turn, the “High development with low PHC and CHW coverage” profile (10.2%) concentrated higher-income municipalities. Despite the favorable economic context, it showed the worst PHC and CHW coverage indicators, suggesting the presence of underserved pockets.

As a validation step, we compared the six profiles with respect to indicators of vaccination status not used in the cluster analysis (Table 3). The spatial distribution followed the profiles: the highest rates of unvaccinated children were concentrated in the North and Northeast, coinciding with the profiles of greater socioeconomic vulnerability and digital exclusion (Figures 4A and 4B). The “High development with operational access barriers”, “High performance in Primary Care”, and “Socioeconomic vulnerability with high operational engagement” profiles showed the highest cumulative IPV and DTP coverage (0-4-year-old cohort) and the lowest values of the VNI. In contrast, the “Socioeconomic vulnerability and digital exclusion” profile concentrated the lowest IPV and DTP coverage, and “High development with low PHC and CHW coverage” showed the highest VNI (3.69), contrasting with the other high-development profiles. Together, these indicators suggest that the classification reflects real differences in the vaccination needs across territories.

**Table 3:** Validation of Municipality Profiles with Vaccination Indicators ^a^.

|  | High performance in Primary Care (N = 1,101) | Socioeconomic vulnerability with high operational engagement (N = 1,394) | Low operational performance (N = 100) | Socioeconomic vulnerability and digital exclusion (N = 955) | High development with operational access barriers (N = 1,188) | High development with low PHC and CHW coverage (N = 536) | Incomplete data (N = 294) |
| --- | --- | --- | --- | --- | --- | --- | --- |
| Inactivated Polio Vaccine (IPV) coverage – dose 3 <sup>b</sup> | 72.1 (65.8 – 77.5) | 69.6 (62.4 - 75.5) | 67.3 (60.1 – 73.6) | 66.4 (60 – 72.9) | 74.5 (68.5 – 79.2) | 68.8 (63.2 – 74.2) | 67.6 (60 – 73.9) |
| DTP coverage - dose 3 (%) <sup>b</sup> | 86.9 (76.4 – 94.6) | 81 (69.5 – 90.4) | 76.5 (66.1 – 90.7) | 75.7 (64.3 – 85.3) | 89.6 (79.8 – 96.6) | 80.7 (70.6 – 89.9) | 79.6 (67.2 – 88.9) |
| MMR coverage - dose 2 (%) <sup>b</sup> | 34.9 (16.3 – 53) | 36.1 (21.2 – 50.2) | 41.8 (27.9 – 54.4) | 35.4 (24.7 – 46.4) | 54.6 (37.8 – 66) | 46 (33.7 – 57) | 40.6 (27.7 – 54.8) |
| Vaccination Needs Index (VNI) <sup>c</sup> | -1.51 (± 4.01) | 0.30 (± 3.81) | 0.71 (± 4.97) | 2.49 (± 3.89) | -2.97 (± 4.24) | 3.69 (± 4.03) | 1.17 (± 5.34) |

Notes:
a) Vaccination coverage data refer to the 0-4 years old cohort.
b) Values expressed as median and interquartile range (IQR).

## DISCUSSION

Our analysis highlights the important role of socioeconomic variables in childhood vaccination in Brazil. Furthermore, the results suggest that the infrastructure and operation of health services exert a strong influence on the outcome, regardless of the social context. Thus, integrating these hierarchical levels is important to understand the complexity of the determinants of childhood immunization in the country.

At the distal level, our findings are consistent with the literature on the social determinants of health [12]. The risk associated with inequality and the protective effect of income and MHDI are consistent with associations reported between socioeconomic conditions and other health outcomes beyond vaccination [36]. The concentration of low vaccination coverage in the North region aligns with historical regional inequalities in Brazil [37].

Under municipal governance, the structural and operational levels have an important impact on the outcome, even after adjustment for socioeconomic variables, consistent with prior evidence linking health service organization to other health indicators [38]. The models obtained illustrate the importance of direct actions in the territory: CHW coverage, keeping UBS open during lunchtime, and the presence of internet in health units were important protective factors. These findings suggest that the reach of PHC is an important component for the success of the PNI.

However, we observed some associations that we considered counterintuitive, for which we propose hypotheses. The risk association of literacy, for example, may reflect its role as a proxy for higher-income and more developed contexts, in which deliberate vaccine refusal tends to be more frequent [39]. Conversely, the associations for the active search for defaulting children and the regular supply of vaccines suggest reverse causality phenomena. Municipalities with lower prior adherence possibly require greater logistical attention and carry out more emergency recovery of defaulters.

It is important to emphasize that the objective of the study was to identify municipal risk profiles, not to estimate the isolated causal effect of each variable. The inclusion of multiple indicators within the same conceptual level may lead to overadjustment, which implies that the coefficients should be interpreted together with the model and not as the total effect of each factor. Although it is difficult to prove causal pathways in ecological studies, we selected the determinants based on the plausibility of their mechanisms, seeking to form more consistent profiles.

Beyond the associations, the cluster analysis showed how several of these factors coexist, suggesting specific profiles of vulnerability and performance. The “High performance in Primary Care” profile stood out with the best structural and operational determinants in the sample, such as high PHC coverage and the percentage of units open during lunchtime. However, this group recorded the worst coverage for the MMR vaccine in the validation step. Unlike IPV and DTP, which are doses of the first year of life, the second dose of MMR is administered later, reflecting a distinct point in the schedule and subject to specific factors that the analyzed determinants may not capture. Therefore, for these municipalities, management should combine maintaining overall service quality with investigating the barriers that may affect MMR vaccination.

Next, the “Socioeconomic vulnerability with high operational engagement” profile showed favorable vaccination performance despite its context of social vulnerability: it recorded higher IPV and DTP coverage and a lower VNI (0.30) than the “Socioeconomic vulnerability and digital exclusion” profile (VNI 2.49), which shares socioeconomic fragilities. This contrast suggests that operational efficiency may partially compensate for distal socioeconomic barriers. For these municipalities, actions should prioritize logistics to facilitate access, as well as avoiding shortages of immunobiologicals.

In contrast, the “Low operational performance” profile showed one of the worst vaccination performances despite having a positive economic context. In this group, we observed a reduction in active search activities and lower CHW coverage. The priority, in these cases, lies in actions with lower financial costs and rapid impact, such as training the local team and establishing monitoring dashboards for recovering defaulters.

The municipalities in the “Socioeconomic vulnerability and digital exclusion” profile concentrated the lowest IPV and DTP coverage. In these territories, the priority is oriented toward investment in infrastructure and human resource training, focusing on computerizing vaccination rooms and information quality. This measure aims at the adequate recording of data and the timeliness of information and enables the prioritized screening of children with delayed schedules.

More favorable socioeconomic contexts also present challenges: municipalities in the “High development with operational access barriers” profile need to adjust staff schedules to keep vaccination rooms open during lunchtime, adapting to the routine of working families or those with difficulties accessing services during business hours. Likewise, “High development with low PHC and CHW coverage” showed lower coverage and higher VNI, indicating that high municipal income alone is not enough. The priority should be expanding PHC and hiring CHW, mapping underserved pockets across the territory, and continuously monitoring service delivery and its quality.

In summary, a favorable socioeconomic context does not guarantee high coverage if health services do not function adequately to meet the population’s access needs. Good operational performance may be decisive and able to overcome more distal barriers. The implications for public health are direct, since the proposed classification works as a guiding diagnostic tool and enables the transition to planning targeted actions, converging with strategies such as the microplanning of high-quality vaccination activities [40].

This study has some limitations that should be considered. First, even though the analyzed outcome corresponds to a time point later than the measurement of the associated factors, and the temporal sequence is plausible, we cannot completely rule out overadjustment resulting from the inclusion of variables at the same level. In addition, the selection of determinants for the clusters required a prior judgment about which associations were intuitive, which introduces an interpretative component into the analysis. The operational indicators, on the other hand, are more subject to temporal variation. Because they were measured before the outcome and may have changed since, we interpret them as characterizing the municipality’s operational context during the study period rather than its current status.

Finally, the consistency and reproducibility of the results depend on the quality of data reporting in the health information systems. It should be noted, however, that the definition of the profiles adopted a conservative criterion. We retained only the associations that were, at the same time, statistically significant and considered intuitive. Although this criterion may exclude real associations whose mechanism we do not understand, it reduces the risk of incorporating spurious findings into the construction of the profiles.

The study has strengths, notably the analysis of almost all municipalities in the country, the use of a hierarchical conceptual model, and the care taken to measure all determinants before the outcome, which is not limited to vaccination coverage but guides another way of analyzing information for decision-making.

As practical implications, the identification of the six municipal profiles and their respective deficiencies in factors associated with vaccination may help formulate more targeted interventions. In profiles where the main problem is operational, it is plausible that strengthening these routines in the territory would generate a positive short-term impact. In those such as “Socioeconomic vulnerability and digital exclusion”, where infrastructure and human-resource qualification problems predominate, investments may show better medium-term effects. Thus, it becomes possible to act on specific vulnerabilities and prioritize resources efficiently, actively contributing to improving the vaccination situation in Brazil.

## CONCLUSION

In this study, we identified multiple contextual factors associated with poliomyelitis vaccination status among children in Brazil. Based on these associations, we classified municipalities into groups that shared similarities in these factors, suggesting common underlying determinants. We expect this classification to guide immunization strategies tailored to the characteristics of each profile, thereby contributing to improved access to vaccination.

## Data Availability

All data produced in the present study are available upon reasonable request to the authors

## SUPPLEMENTARY MATERIAL

### Full list of questions used for the structural and operational factors

- Does the BHU* monitor up-to-date vaccination of pregnant women?
- Does the BHU monitor the vaccination status of children up to 2 years of age?
- Does the BHU conduct active outreach/searches for children with delayed vaccination?
- BHU conduct active outreach/searches for children (regardless of the reason)?
- Is there a Community Health Worker (CHW) on the team?
- Is there a population not covered by CHWs?
- Does the BHU provide the community with a list of the services it offers?
- Does the BHU open at lunch hours?
- Does the BHU have a dedicated vaccination room?
- Does the BHU have internet access?
- Does the BHU always have printed vaccination cards available?
- Does the BHU provide vaccination?
- Does the BHU regularly offer vaccines?
- Does the BHU always have theTdap vaccine available?
- Does the BHU always have the Oral Polio Vaccine (OPV) available?
- Does the BHU always have the Inactivated Polio Vaccine (IPV) available?
- Does the BHU always have the Measles, Mumps, and Rubella (MMR) vaccine available?
- Does the BHU always have the Pentavalent vaccine available?

**\*BHU: Basic Health Unit**

**Table S1:** Descriptive statistics and correlation matrix of childhood vaccination coverages (Brazil, 2024)

| Vaccine | Mean coverage (%) | Standard Deviation (%) | Inactivated Polio (IPV) <sup>a</sup> | DTP / Pentavalent <sup>a</sup> | MMR <sup>a</sup> |
| --- | --- | --- | --- | --- | --- |
| Inactivated Polio (IPV) – 3rd dose | 93.56 | 9.21 | 1.00 |  |  |
| DTP / Pentavalent – 3rd dose | 93.70 | 9.24 | 0.98 <sup>b</sup> | 1.00 |  |
| MMR (Measles, Mumps, Rubella) | 87.09 | 14.83 | 0.61 <sup>b</sup> | 0.60 <sup>b</sup> | 1.00 |

Notes:
a) Correlation values represent Pearson's correlation coefficient.
b) $p < 0.001$

**Table S2:** Hierarchical Poisson regression models with associated factors with the rate of unvaccinated children (IPV Vaccine, third dose, 2024)

| Standardized (Z-score) variables | Model 1 (Socioeconomic) | Model 2 (Health services structure) | Model 3 (Operational) |
| --- | --- | --- | --- |
| *Level 1: Socioeconomic* | RR (CI 95%) <sup>a</sup> | RR (CI 95%) <sup>a</sup> | RR (CI 95%) <sup>a</sup> |
| Gini index | **1.33 (1.32 – 1.34)** | 1.26 (1.25 – 1.27) | 1.24 (1.24 – 1.25) |
| Life expectancy, in years | **0.93 (0.92 – 0.94)** | 0.95 (0.94 – 0.96) | 0.95 (0.94 – 0.96) |
| MDHI | **0.61 (0.60 – 0.62)** | 0.77 (0.76 – 0.78) | 0.77 (0.76 – 0.79) |
| Rural population | **0.71 (0.71 – 0.72)** | 0.74 (0.74 – 0.75) | 0.76 (0.75 – 0.77) |
| PCI (R\$) | **0.98 (0.98 – 0.99)** | 0.96 (0.95 – 0.97) | 0.96 (0.95 – 0.97) |
| Literacy rate | **1.73 (1.71 – 1.75)** | 1.52 (1.50 – 1.53) | 1.47 (1.45 – 1.49) |
| Child work | **0.94 (0.93 – 0.95)** | 1.01 (1.00 – 1.02) | 1.01 (1.00 – 1.02) * |
| *Level 2: Health services structure <sup>c</sup>* |  |  |  |
| PHC coverage | - | **0.88 (0.88 – 0.89)** | 0.90 (0.89 – 0.90) |
| Internet on vaccine room | - | **0.82 (0.82 – 0.82)** | 0.83 (0.82 – 0.83) |
| Computers on vaccine room | - | **1.00 (0.99 – 1.00)** | 1.00 (1.00 – 1.00) |
| *Level 3: Operational <sup>c</sup>* |  |  |  |
| Always offers vaccine | - | - | **1.05 (1.04 – 1.05)** |
| Opens in lunch hours | - | - | **0.98 (0.97 – 0.98)** |
| Does active search | - | - | **1.02 (1.01 – 1.03)** |
| Does not perform active search | - | - | **1.02 (1.01 – 1.03)** |
| Population without CHWs coverage | - | - | **1.15 (1.14 – 1.15)** |

Notes:
a) RR: Rate Ratio. 95% CI: 95% Confidence Interval. Standardized variables (Z-Scores) for the Poisson analysis.
b) The measures of association reflect the hierarchical approach, presenting total effects for each level, with levels 2 and 3 adjusted by its predecessors.
c) Model 1: Total effects of the distal (socioeconomic) level. Model 2: Total effects of the intermediate (structure) level and direct effects of the distal level, not mediated by intermediate. Model 3 (fully adjusted): Total effects of the proximal (operational) level, adjusted for the preceding levels and direct effects of the distal and intermediate levels not mediated by operational factors.
\* p = 0.15. All other measures of association presented in this table had p < 0.01

